# Changing hand appearance using visual illusions modulates body perception disturbance and pain in longstanding Complex Regional Pain Syndrome: A randomised trial

**DOI:** 10.1101/2020.02.26.20020420

**Authors:** Jennifer S Lewis, Roger Newport, Gordon Taylor, Mike Smith, Candida S McCabe

## Abstract

Effective treatment of longstanding Complex Regional Pain Syndrome (CRPS) is a challenge, as causal mechanisms remain elusive. People with CRPS frequently report distorted subjective perceptions of their affected limb. Evidence of pain reduction when the affected limb is visually altered in size, suggests that visual illusions used to target central processing could restore coherence of this disrupted limb representation. We hypothesised that using virtual reality that alters hand image to match the patient’s desired hand appearance, would improve body perception disturbance and pain. Also, repeated exposure would maintain any therapeutic effect.

A blinded randomised controlled trial of 45 participants with refractory upper-limb CRPS and body perception disturbance (BPD) viewed a digital image of their affected hand for one minute. The image was digitally altered according to the patient’s description of how they desired their hand to look in the experimental group and unaltered in the control group. BPD and pain were measured pre and post-intervention. A subgroup was followed up two weeks after a course of repeated interventions.

BPD (p=0.036, effect size (ES)=0.6) and pain intensity (p = 0.047, ES=0.5) reduced in 23 participants after single exposure compared to controls (n=22). At follow-up the subgroup (experimental n= 21; control n=18) showed sustained pain reduction only (p=0.037 ES=0.7), with an overall 1.2 decrease on an 11-point scale.

Visually changing the CRPS hand to a desired appearance modulates BPD and pain suggesting therapeutic potential for those with refractory CRPS. Future detailed studies to optimise this therapeutic effect are required.

## INTRODUCTION

The recalcitrance of longstanding Complex Regional Pain Syndrome (CRPS) to treatment is a challenge for the international pain community. Whilst most CRPS cases recover within the first twelve months, 27% of patients suffer from persistent symptoms that develop into a long-term condition [1]. In these cases, CRPS symptomatology considerably impacts on function, emotional and social wellbeing and poses an economic burden on society [1,2,9].

Successful treatment of longstanding CRPS remains problematic as therapeutic responses to conventional pharmacological options are limited[13], since the underlying mechanisms of CRPS are still unknown. As such, therapeutic targets remain elusive [5]. There is little evidence to support the gold standard treatment of multidisciplinary rehabilitation which is costly and resource-intensive [22,37,38,40]. To address this challenge, it is crucial to develop approaches that are both clinically and cost effective.

People with CRPS frequently report distorted subjective perceptions of their affected limb. These can manifest as perceptual changes in affected limb size and shape, a dislike in appearance and a loss of ownership of their painful limb [14,17,26]. Body perception disturbances comprise both alterations in sensorimotor representation e.g. perceived changes in size and shape (body schema) and a perceptual awareness of the limb e.g. dislike of appearance and disownership of the limb (body image) [31]. Evidence suggests that these body perception disturbances are associated with maladaptive cortical representation of the limb [21,32]. Body perception disturbance has been shown to positively correlate with CRPS pain intensity [15].

Novel drug-free technologies such as virtual reality have revealed analgesic effects in acute and chronic pain states [7,34]. Specifically, the use of body illusions to relieve clinical pain shows therapeutic promise [3]. Body representation is highly adaptive, as various illusions that change the shape of the painful body have shown [10, 35]. These illusions alter central body representation– multiple dynamic multisensory maps of the body that are constantly updated by somatosensory, visual, proprioceptive, vestibular inputs and motor feedback [20]. That this sense of the bodily self persists even when sensory input stops, emphasises the robustness of our body representation [23].

Given that central mechanisms are the primary driver in persistent pain [39], manipulating central systems as a potential target in the treatment of pain, seems a logical route for exploration [30]. Therefore, we propose using body illusions in CRPS to address body perception disturbance in order to target centrally-mediated maladaptive body representations, which may in turn, reduce pain.

Using Mediated Virtual Reality (MVR) in longstanding CRPS, we aim to alter the appearance of the affected hand based on how those with CRPS would like their hand to look. We postulate that a match between the visual appearance and desired representation of the CRPS affected hand, would normalise the maladapted central representation of the hand. We hypothesise that a visual illusion to improve the subjective appearance of the affected hand would (1) normalise body perception, ownership and liking of the hand, which would lead to (2), a reduction of pain, and (3) sustain a therapeutic effect with repeated exposure for those with longstanding CRPS.

## METHODS

### Participant recruitment and group assignment

Potential participants were identified from the CRPS UK network registry (crpsnetworkuk.org/Registry.php) and clinics at The Royal National Hospital for Rheumatic Diseases, Royal United Hospitals Bath NHS Foundation Trust, Bath and The Walton Centre NHS Foundation Trust, Liverpool, UK. Those who met the following study inclusion criteria were recruited for this convenience sample; met the Budapest clinical diagnostic criteria [12] for CRPS affecting one upper limb; aged 18 and over and had no co-morbidity that might influence CRPS symptoms i.e. stroke, diabetes and fibromyalgia.

The sample size for this study was based on MIRAGE illusion within-subject pilot data in 14 CRPS participants. A total sample size of 88 participants (44 per group) was calculated as sufficient for a mean (SD) reduction in the pain numerical rating scale (primary outcome) of 1.733 (2.89) points, on a 0-10 scale with a power of 80% and a 0.05 two-sided significance.

Participants gave written informed consent prior to participation in procedures approved by local hospital and University ethics committees in accordance with the Declaration of Helsinki. The registered ISRCTN trial number is ISRCTN64093359 (www.isrctn.com/ISRCTN64093359). Following informed consent and to avoid selection bias, participants were randomly allocated to either the manipulation (experimental) or non-manipulation (control) group. In advance of testing, a person independent of data collection used a computer generated random sequence to produce information regarding group allocation that was placed in sealed and numbered envelopes. Envelopes were sequentially opened by the MIRAGE operator after consent and prior to testing. To minimise performance and detection bias, the clinical assessor and participants were blinded to group allocation. Participants were not informed of the study hypothesis to minimise responder bias.

### Experimental Procedure

Participants attended up to five sessions comprising four intervention sessions and a final follow-up session two weeks later. A detailed schematic of the study procedure is illustrated in Figure 1. Data were collected within laboratory-controlled conditions. The primary outcome measures collected were metrics directly related to our hypotheses (Body perception disturbance, pain intensity, perceptual ratings).

**Figure 1:**
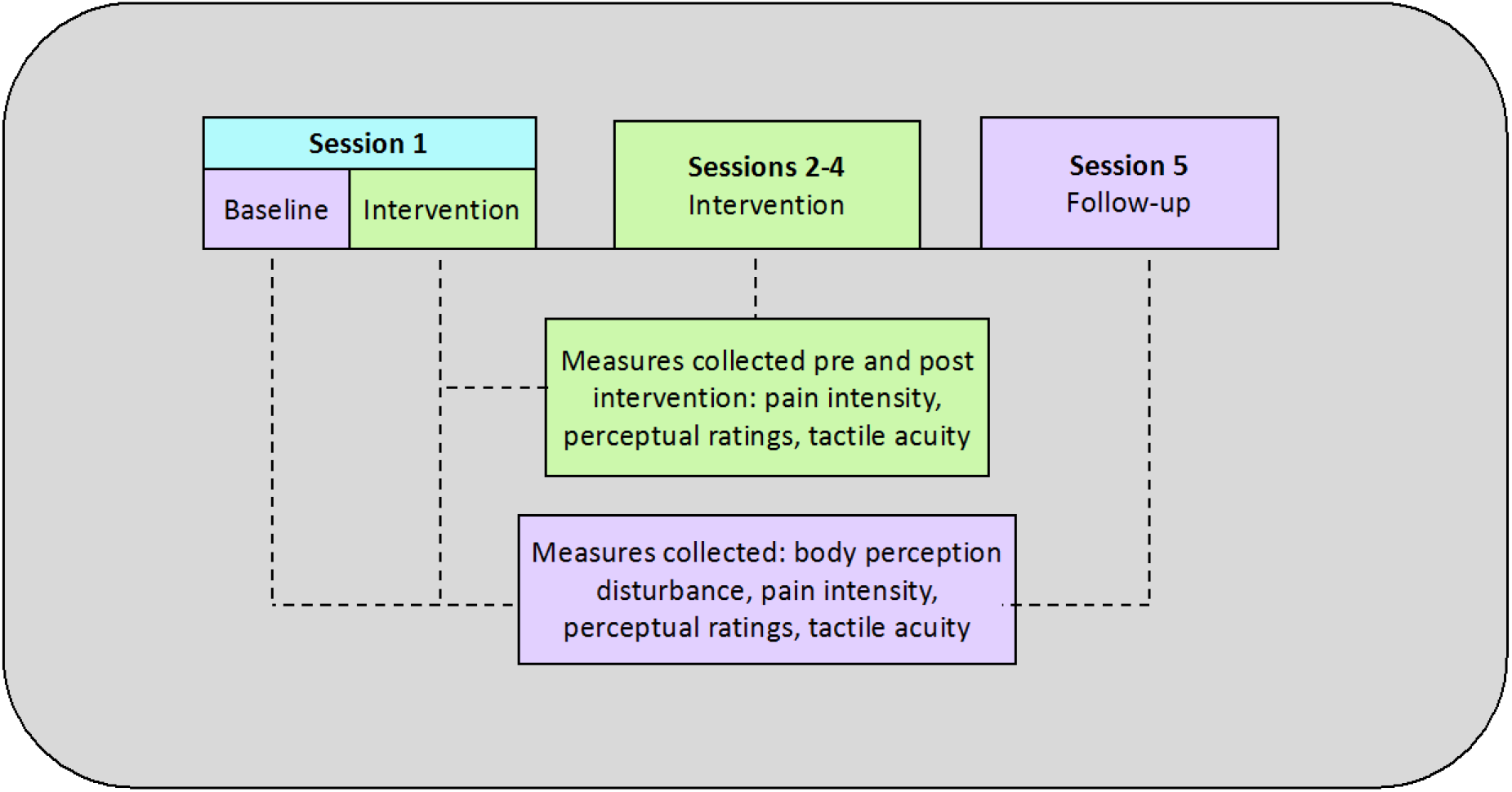
Study procedure.

Intervention sessions (sessions 1-4) consisted of three parts: (A) pre-intervention, (B) intervention, and (C) post-intervention. The intervention using the MIRAGE system [35], a non-invasive MVR device, consisted of either visual illusions involving the digital manipulation of the appearance of participants’ hands for the manipulation group (MG) or non-manipulation for the non-manipulation group (NG). Baseline primary and secondary outcome measures were collected prior to the initial intervention within the first session (see Figure 1).

#### Baseline (conducted outside the MIRAGE system)

In a seated position, participants directly viewed their actual hands positioned palm down on a table at waist height in front of them and outside of the MIRAGE system. Primary and secondary baseline measures were collected. For purposes of describing pain characteristics participants also completed The Neuropathic Pain Symptom Inventory (NPSI) [4]. The NPSI is a validated measure for the severity of neuropathic pain. The questionnaire determines subjective intensities (for the preceding 24 hours) of spontaneous superficial, spontaneous deep, paroxysmal and evoked pain, as well as paraesthesia. These different neuropathic symptoms are rated on an 11-point numerical rating scale. A total score is calculated by summing the five categories. Higher scores denote greater intensity.

##### (A) Pre-intervention (conducted within the MIRAGE system)

In a similar seated position to that at baseline, participants sat with each arm placed into one of the two apertures of the MIRAGE system so that both hands rested palm down on a flat surface within the system. Participants viewed a real-time digital video image of their hands through a horizontal ‘window-like’ surface above and perpendicular to these apertures. The image of their hands was displayed via this surface in such a way that their hand image appeared to be in the same physical and spatial location as their actual hands. The participant viewed their affected hand image within the device prior to the intervention. Primary outcome measures were collected as described below.

##### (B) Intervention (conducted within the MIRAGE system)

###### i) Manipulation

As the participant viewed their affected hand within the device, the MIRAGE system operator digitally altered the appearance of the painful hand using specifically designed software via a laptop (MacBook Pro 15” Model ME664B/A using Windows 7 running LabView 2012 (National Instruments), as part of the MIRAGE system. In response to the specific description given by each participant, changes were made in real time to aspects of shape, size and/or colour of the hand, based on how they wished their hand to look i.e. their desired hand appearance. Participants rated their satisfaction of hand appearance whilst looking at the hand image by answering the question “*How satisfied are you with the hand as you see it?”* on a 7-point Likert scale ranging from −3 (strongly dissatisfied) to +3 (strongly satisfied). If participants rated <+1, the image was further altered to reach a rating of +1 in order to better match the participant’s desired hand appearance. Requests were specific to the individual, therefore resulting hand images were unique on each occasion and took up to a minute to complete.

Once they were satisfied, participants viewed the resultant image for one minute. No visual changes were made to the unaffected hand. Post-intervention measures were collected following this procedure.

###### ii) Non-manipulation

The procedure and duration for non-manipulation was exactly the same as that described in i) manipulated condition by the operator appearing to click the computer keys with the exception that the image was not actually visually altered, though the participant believed it to have been. A satisfaction level of <+1 was not required in the control group in order to proceed to the intervention. The hand image was viewed for one minute and followed by post-intervention data collection.

##### (C) Post-intervention (conducted within the MIRAGE system)

The same measures taken at pre-intervention were repeated post-intervention.

#### Primary Outcome Measures

Primary outcomes were measured at baseline, pre-intervention and post-intervention as follows:

##### a. The Bath body perception disturbance (BPD) scale

The BPD scale was used to measure changes in body perception of the affected limb [14]. This scale demonstrates good internal consistency and adequate interrater reliability in a CRPS sample [15] A higher score indicates a greater degree of disturbance (see [14] for a description of the scale).

##### b. Pain intensity numerical rating scale (NRS)

To assess current pain, participants verbally rated their affected hand pain intensity on an 11-point NRS anchored at 0 (no pain) and 10 (worst pain imaginable).

##### c. Perceptual statement ratings

Perceptual statement ratings were adapted from Schaefer et al. [36]. These were used to assess subjective perceptual changes associated with the affected hand. Whilst viewing their hand, participants provided a verbal rating to the following statements on a 7-point Likert scale ranging from −3 (strongly disagree) to +3 (strongly agree): *(1) It feels like the hand that I am looking at is my hand; (2) I like the appearance of my hand as I see it; (3) I feel my hand is lighter; (4) I feel my hand is heavier; (5) I feel my hand is different in sensation*.

### Statistical analyses

To test our hypotheses that a visual illusion to improve the subjective appearance of the affected hand would (1) normalise body perception, ownership and liking of the hand, and (2) reduce pain, we calculated post intervention changes in these measures at session 1 by subtracting the respective pre-intervention score from the post-intervention score. Parametric tests (independent t-tests) were performed on group mean change scores to compare pre-post intervention change in outcome measures between the two groups (MG and NG) (n=45).

Our third hypothesis, that repeated exposure would sustain a therapeutic effect, was tested with a repeated measures ANOVA with a Greenhouse-Geisser correction, on participants (n=39) that completed five sessions. An independent t-test was performed to explore changes in pain at baseline (session 1) when compared to follow-up (session 5) between these two groups. Statistical significance levels were set at p=0.05. Effect sizes for each comparison were calculated by dividing the mean difference between groups by the pooled standard deviation. Confidence intervals were calculated at 95%. All analyses were undertaken using IBM SPSS Statistics v.23 (IBM corp. Armonk, NY USA.

## Results

A total of 46 participants were assessed for eligibility (Figure 2). One patient did not meet the Budapest clinical criteria for CRPS [12] of one arm on examination and was excluded. Forty-five participants [29 females, aged (mean ±SD) 52 ±13 years, mean disease duration 56 ±54 months (4.7 years)] were randomised to either the manipulation (experimental) group (n=23) or the non-manipulation (control) group (n=22) and completed session 1. Individual participant characteristics are presented in supplemental data (SD) (Table i). The total sample did not reach the expected sample size as Registry administrators fed back that potential participants felt unable to tolerate travelling to multiple sessions due to persistent pain.

**Figure 2.**
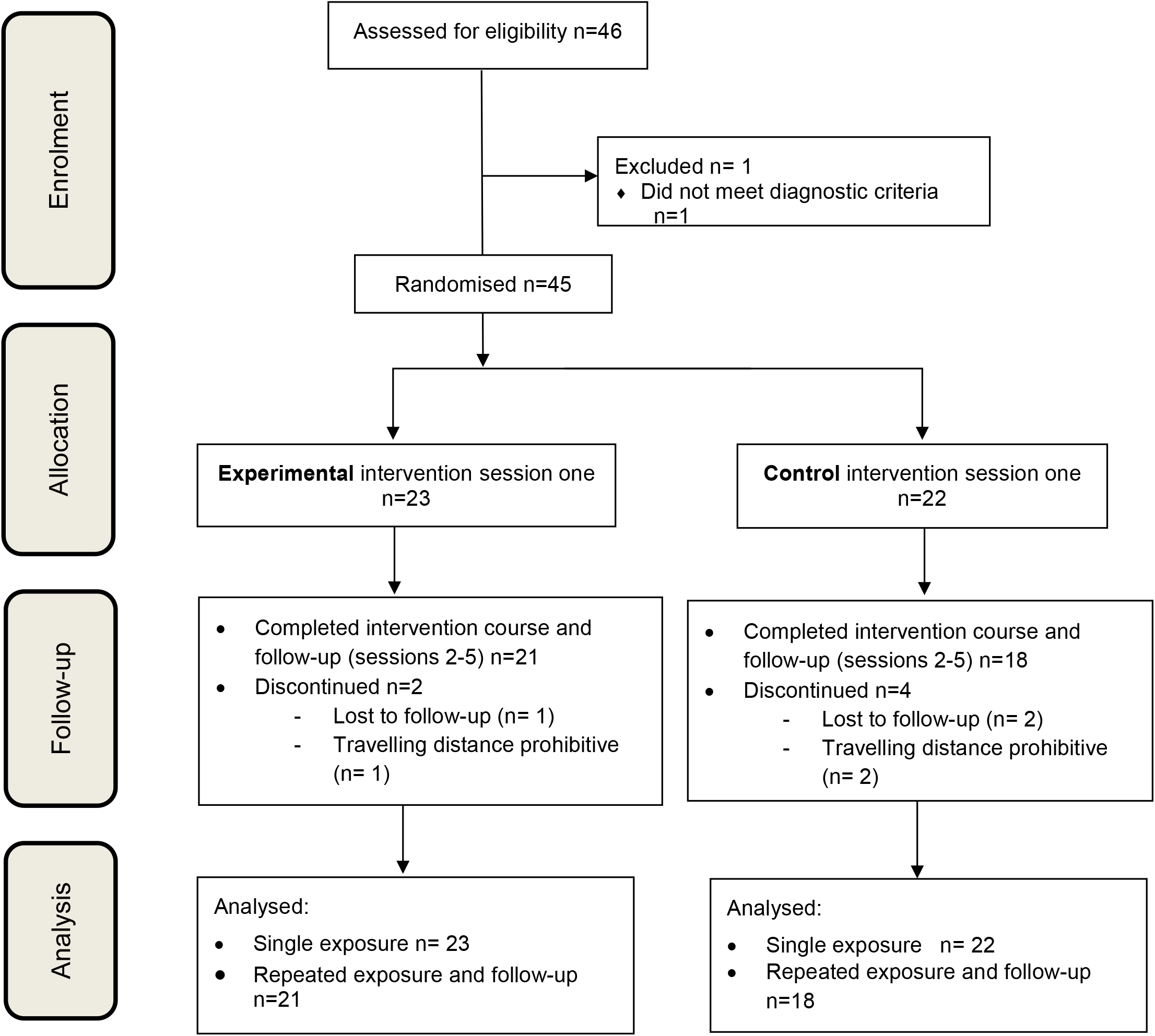
CONSORT flow diagram.

Twenty-one of the Manipulation Group (MG) completed a course of four intervention sessions and a follow-up, whilst 18 of the Non-Manipulation (NG) group completed the intervention course and follow-up. Analysis of data was conducted in 45 participants for single exposure and in 39 participants for repeated exposure and follow up (Figure 2). There were no significant differences in demographic and clinical characteristics between the two groups at baseline (Table 1)

**Table 1:**
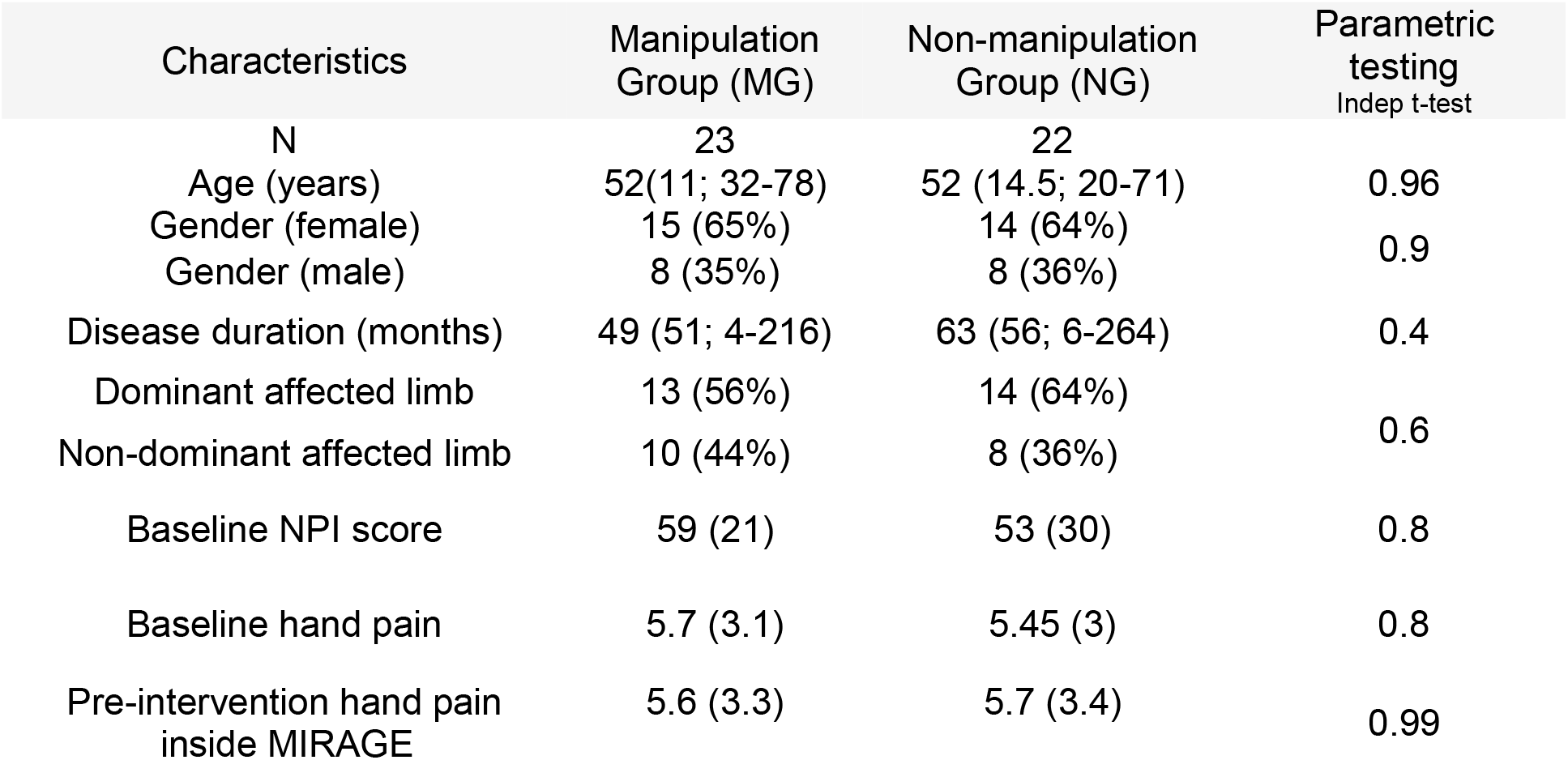
Participant demographics and clinical characteristics.

Participants were very specific about how and the degree to which changes to hand appearance were made (i.e. lengthening fingers or narrowing the dorsum of the hand). Reshaping (enlarging or reducing) of precise areas such as individual digits was considered important by the participant to achieve their desired appearance. These hand images were individual to the participant and unique to each study session.

### Single exposure (Hypotheses 1&2)

Body perception disturbance: A significant reduction in the Bath BPD scale total score (t=2.16, df=43, p=0.036) for pre-post intervention differences in MG (mean, ±SD) (−6, 7.9) was found when compared to NG (−1.3, 6.5). See Figure 3A. Effect size (ES)= 0.64 (0.042,1.25).

**Figure 3:**
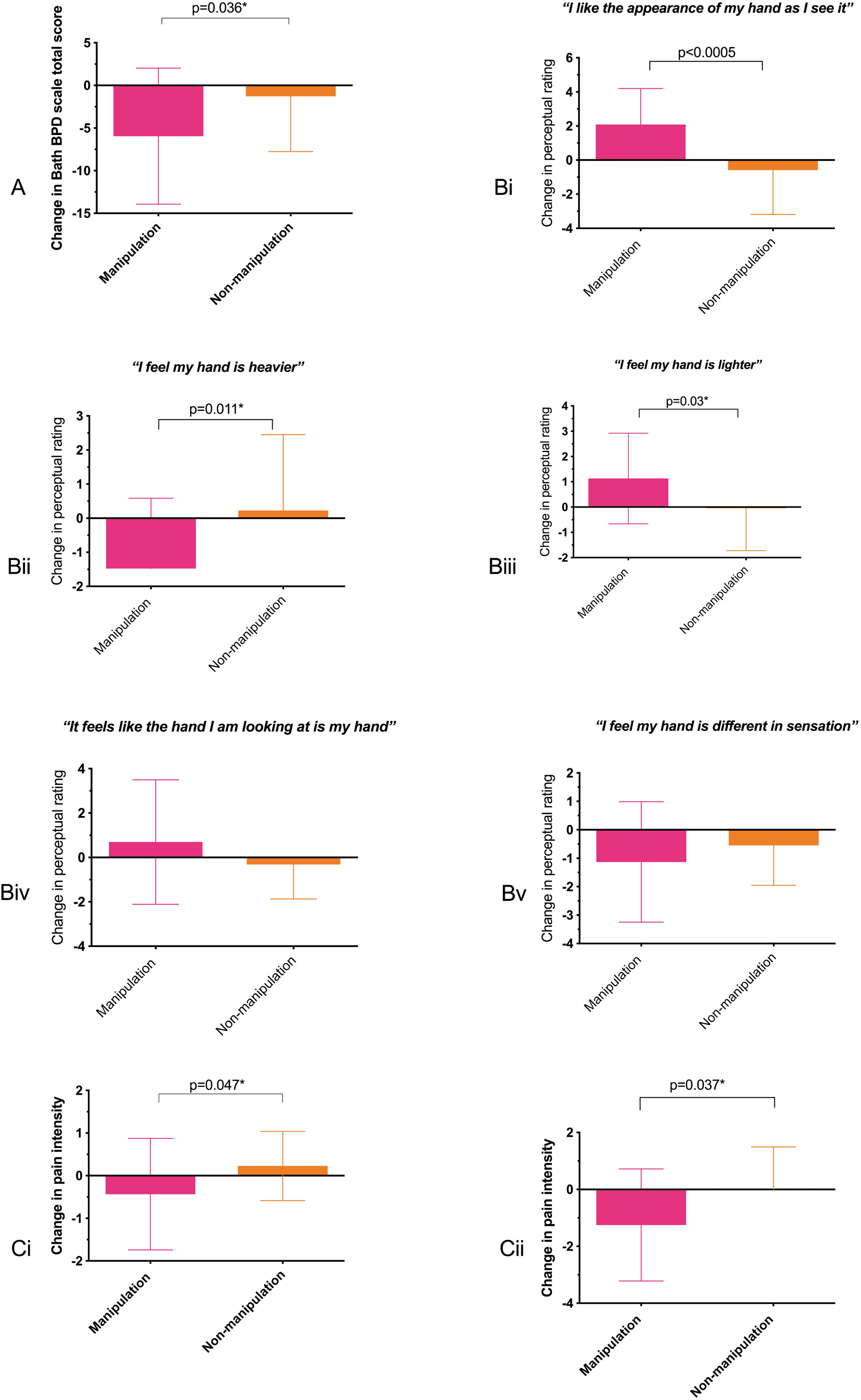
A. Body perception disturbance:single exposure. B i-v Perceptual ratings:single exposure C. Pain: i.single exposure ii.repeated exposure.

Perceptual ratings: A significant difference (t=3.81, df=43, p<0.0001) in pre-post intervention changes between MG (2.1, 2.1) and NG (−0.6, 2.6) for perceived liking of the affected hand represented an improvement in liking of hand appearance, ES= 1.135 (−1.73, −0.53). Sense of heaviness was significantly reduced (t=2.67, df=43, p=0.011) post intervention for MG (−1.5, 2) compared to NG (0.23, 2), ES= 0.8 (0.193,1.39). A significant difference (t=2.27, df=43, p=0.03) in pre-post intervention changes for perceived lightness was found between MG (1.24,1.8) and NG (−0.05, 1.7), ES= −0.7 (−1.28, −0.07). Taking the change in rating of perceived lightness and heaviness together, demonstrates an overall perception that the hand felt lighter post intervention. No pre-post intervention differences were found between the groups for ownership (t=1.49, df=43, p=0.14), ES= -−0.45 (−1.05, 0.16) and sensation (t=1.09, df=43, p=0.28), ES= 0.32 (−0.28,0.93). Perceptual rating results are presented in Figure 3Bi-v. (See SD Table ii for mean perceptual rating scores pre /post intervention).

Current pain intensity: A significant reduction in current pain (t=2.03, df=43, p=0.047) was found between pre-post intervention pain differences for MG (−0.43, 1.3) compared with NG (0.23, 0.8) (Figure 3Ci), ES = 0.46 (0.003, 0.92).

In summary, we found that a single one-minute exposure to an illusion that visually altered affected hand appearance to a desired look, significantly reduced body perception disturbance, improved liking and increased perceived lightness of the affected hand. There was also a significant decrease in pain.

### Repeated exposure (Hypothesis 3)

#### Pain

A repeated measures ANOVA of mean pre-post intervention changes in pain (sessions 1-4) revealed a significant effect of repeated illusory exposure on pain reduction in MG F(3,105)=1.89, p=0.014) when compared to NG.

Furthermore, to establish whether the effects of illusory exposure were sustained for two weeks after repeated exposure, the mean changes for pain intensity at baseline (session 1) and follow-up (session 5) were compared between the groups. This revealed a significant reduction in pain intensity in MG (t=2.18, df=36, p=0.037) when compared to NG showing an overall mean pain reduction in MG of 1.2 on an 11-point scale at follow-up (MG mean= −1.19, NG mean=0) (Figure 3Cii), ES = 0.7 (0.05,1.38).

#### Body perception disturbance and perceptual ratings

When comparing measures at baseline (session 1) to follow-up (session 5) there were no between group differences in the Bath BPD scale (t=-0.03, df=37, p=0.98), ES= −8.8 (−0.65, 0.63) perceptual ratings of ownership (t=0.495, df=37.8, p=0.62), ES= 0.16 (−0.49, 0.8) liking of appearance (t=0.62,df=37,p=0.53), ES= 0.2 (−4.7, 8.9) lightness (t=1.8,df=38, p=0.08), ES= 0.6 (−1.4, 2.8) heaviness (t=0.7, df=34, p=0.5), ES= 0.22 (−3.6, 7.3) or difference in sensation (t=0.6, df=39,p=0.6), ES= 0.12 (−4.05, 7.5).

## DISCUSSION

Findings confirm our hypotheses that short exposure to a visual illusion which matches the desired appearance of the painful hand in longstanding CRPS, 1) normalises body perception disturbance, 2) reduces pain and 3) sustains a therapeutic effect with repeated illusory exposure. However, there was no effect on perceived ownership of the hand. To aid interpretation, we present these findings within the context of a conceptual model.

Our results are consistent with Pitron et al’s model of body representation [33]. This ‘*serial co-construction*’ model enhances previous conceptual models of body representation in clinical conditions by presenting body schema and body image as two distinct yet interacting concepts whereby body schema modifies body image [25,28]. Responses to the illusion were rapid and resulted in changes to a range of sensory-perceptual and cognitive experiences. This suggests that illusory exposure rapidly modifies the more malleable body image by visually presenting a perceptually more appealing affected hand that triggers these changes.

How the more enduring configuration of body schema interacts is of particular interest [18,33]. That participants were specific about how and to what degree they wished their hand appearance to change, indicates access to a vividly preserved yet dormant pre-CRPS hand representation. This is potentially important as it further demonstrates that different body representations co-exist and can be accessed in the brain. We deduce that participants spontaneously accessed their longstanding body schema. The concept of an inherent body schema is reminiscent of children with congenital aplasia who experience a phantom limb suggesting the presence of an innate cortical limb representation [23].

Reshaping of specific areas was considered important by the participant to achieve the desired appearance reflecting their unique body schema map. Individualised virtual reshaping offers potential for an advanced approach to treatment from that of uniformly resizing the whole hand as previous CRPS studies have done [32].

The interplay between the desired hand image and perceptual responses reflects the interaction between body image and body schema. That CRPS presents with simultaneous deficits in body schema and image is similar to eating disorders where evidence of distortions in these two distinct types of body representation have been shown [25].

So how might viewing the desired appearance of the painful hand ameliorate pain? We propose that the visually desired hand stimulus matches that of the innate body schema triggering a congruence to be restored. Short illusory exposure rapidly reduced pain in people with longstanding CRPS, representative of the 27% with treatment resistant chronic disease [1]. Reconfiguration between cognitive and perceptual representations of body image with the innate body schema resumes congruent multisensory processing and could explain why participants express a restorative change in hand perception. Pain may result from an incongruence between body image and body schema, hence once congruence is restored pain rapidly ameliorates. Indirectly modulating pain by directly targeting body perception disturbance supports a relationship between body perception disturbance and pain [15].

In CRPS the innate body schema may be suppressed by pain such that the altered hand representation which CRPS patients describe, becomes dominant. Pain related body perception disturbances may be caused by maladaptive cortical plasticity that is continually maintained by this incongruence so preventing the innate body schema from being restored to the ‘working’ schema. Perhaps this explains why it is a challenge to therapeutically correct [16]. Based on our interpretation, we propose an updated conceptual body perception disturbance model to that of Pitron et al.[33] (Figure 4).

**Figure 4.**
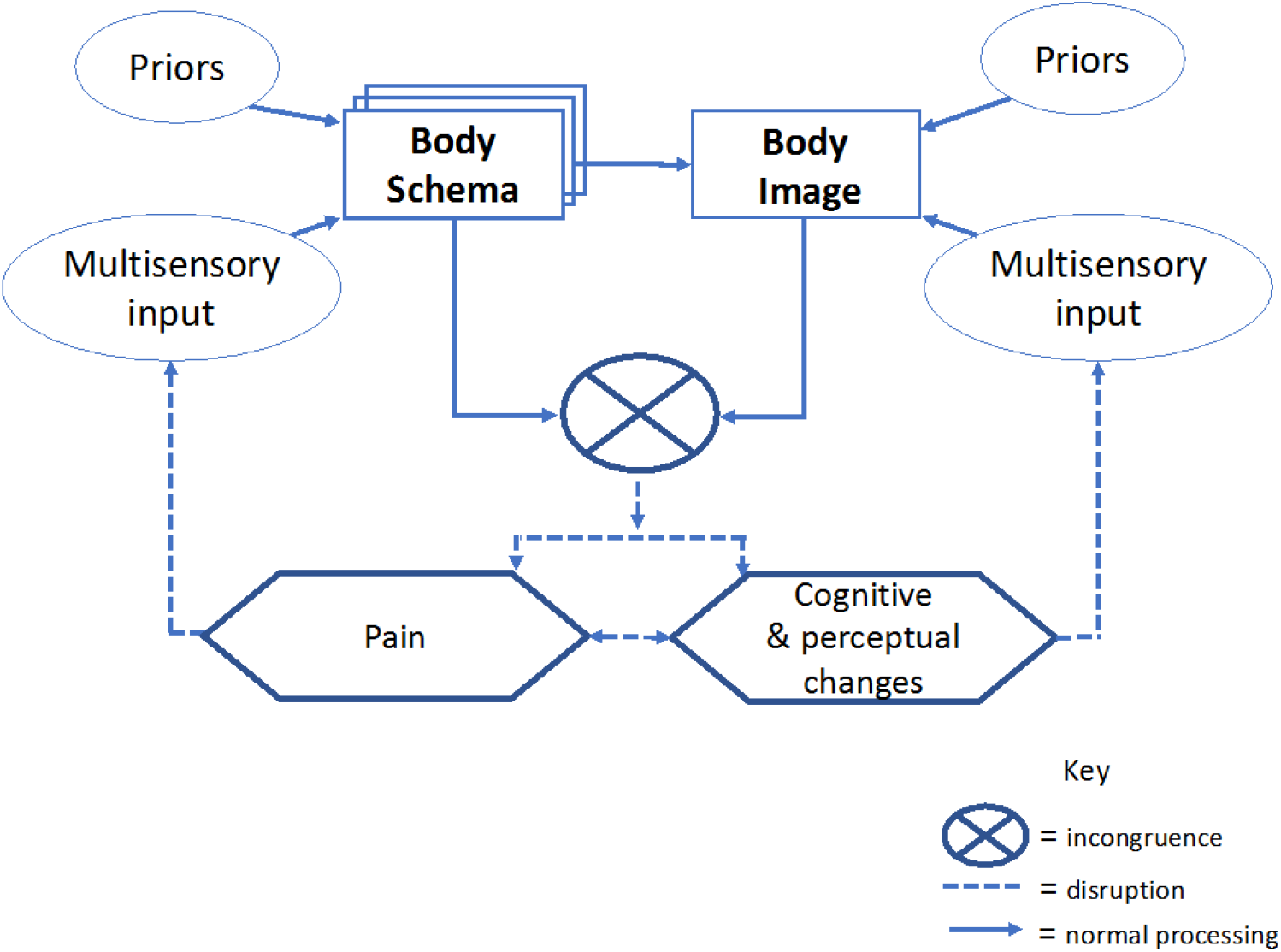
Body representation co-construction model modified from Pitron et al. [33].

Neuroimaging studies of body representation processing show activation in the posterior parietal cortex [6]. Furthermore, our results support previous findings that visual manipulation triggers alterations in body self-perception demonstrating that visual inputs involving the occipital cortex influence higher order multisensory processing associated with body representation [36]. The lateral (extrastriate body area) and medial (fusiform body area) occipitotemporal cortices may also be involved given associations with visual representation of body shape. [8]. Future brain imaging studies of real-time exposure to body reshaping illusions would add valuable insight into neural mechanisms.

Findings concur with Peltz et al. [32] where a positive correlation was revealed between an overestimation of hand size and increased inattention/neglect-like feelings-a potential feature of body perception disturbance. Our findings suggest that a more desirable-looking hand improves attention to the hand as disturbances in body perception are reduced. However, it is difficult to draw a conclusion regarding the contribution of inattention/neglect to body perception disturbance as contradictory results from other studies have found no association between neglect and pain [24].

Interestingly, pain relief was maintained for up to two weeks after repeated illusory exposure providing a sustained therapeutic effect for those with refractory CRPS. Unlike pain however, changes in body perception disturbance were not sustained at follow-up. The frequency of these short illusions over a month was perhaps insufficiently powerful to maintain an effect.

Ownership, considered part of body image [19], remained unchanged following either single or repeated exposure. This is contrary to our conceptual model (Figure 4) and previous results where experimental manipulation of the upper limb produced changes in ownership [27]. Perhaps the illusion was of insufficient strength and/or duration to restore ownership.

## Data Availability

The data is the copyright of the authors. Permission is required to use the data

## Limitations

The sample size is smaller than expected which reflects the difficulty in undertaking complex intervention studies within a chronic pain population. Although the illusion reduced pain, we note that our results do not meet the clinical significance threshold of a two-point reduction in NRS [11]. However, these results are encouraging and suggest further work is required to maximise the effect and reach clinical significance. Also, the period between repeated exposure and follow-up was short therefore, how long the therapeutic effect might last is unknown.

Further to previous pain-relieving treatments that evoke a virtual limb [29], our findings suggest the treatment potential of a ‘desired-appearance illusion’ for patients previously considered to have refractory disease. Future work is required to explore the optimum duration and frequency of the illusion for best therapeutic effect. Additionally, how the intervention can be adapted for suitable delivery in a clinical setting could be established.

In summary, we found in patients with refractory disease, that single exposure to a visual image of the CRPS hand, digitally manipulated to match the subjective desired hand appearance, has a therapeutic effect on body perception disturbance and pain. Repeated illusory exposure sustained this effect in pain. A reduction in body perception disturbance and pain in patients with refractory disease suggests exciting treatment potential for CRPS and other chronic pain conditions where body perception disturbances arise. Future studies are required to determine dosage and clinical suitability in order to achieve sustained relief for those with longstanding CRPS.

## Acknowledgements

Sincere thanks go to all the patients that freely gave their time to participate in this study. We would like to thank the clinical and research staff responsible for pain research at the Royal National Hospital for Rheumatic Diseases, Bath and the Walton Centre in Liverpool for their dedication in identifying potential participants. My gratitude goes to Dr Massih Moayedi for his advice and comments. This paper presents independent research funded by the National Institute for Health Research (NIHR). The views expressed are those of the authors and not necessarily those of the NHS, the NIHR or the Department of Health.

## References

[1] Bean DJ, Johnson MH, Heiss-Dunlop W, Kydd RR. Extent of recovery in the first 12 months of complex regional pain syndrome type-1: A prospective study. European Journal of Pain 2016;20:884–894.

[2] Bean DJ, Johnson MH, Heiss-Dunlop W, Kydd RR. Factors Associated With Disability and Sick Leave in Early Complex Regional Pain Syndrome Type-1. Clin J Pain 2016;32:130–138.

[3] Boesch E, Bellan V, Moseley GL, Stanton TR. The effect of bodily illusions on clinical pain: a systematic review and meta-analysis. Pain 2016;157:516–29.

[4] Bouhassira D, Attal N, Fermanian J, Alchaar H, Gautron M, Masquelier E, Rostaing S, Lanteri-Minet M, Collin E, Grisart J, Boureau F. Development and validation of the neuropathic pain symptom inventory. Pain 2004;108:248–257.

[5] Bruehl S. Complex regional pain syndrome. BMJ-British Medical Journal 2015;351:h2730.

[6] Buccino G, Binkofski F, Fink GR, Fadiga L, Fogassi L, Gallese V, Seitz RJ, Zilles K, Rizzolatti G, Freund H-. Action observation activates premotor and parietal areas in a somatotopic manner: An fMRI study. Eur J Neurosci 2001;13:400–404.

[7] Chan E, Foster S, Sambell R, Leong P. Clinical efficacy of virtual reality for acute procedural pain management: A systematic review and meta-analysis. PLoS ONE 2018;13.

[8] Costantini M, Urgesi C, Galati G, Romani GL, Aglioti SM. Haptic perception and body representation in lateral and medial occipito-temporal cortices. Neuropsychologia 2011;49:821–829.

[9] de Mos M, de Bruijn, A G J, Huygen, F J P M, Dieleman JP, Stricker BHC, Sturkenboom, M C J M. The incidence of complex regional pain syndrome: A population-based study. Pain 2007;129:12–20.

[10] Diers M, Zieglgaensberger W, Trojan J, Drevensek AM, Erhardt-Raum G, Flor H. Site-specific visual feedback reduces pain perception. Pain 2013;154:890–896.

[11] Farrar JT, Young JP, LaMoreaux L, Werth JL, Poole RM. Clinical importance of changes in chronic pain intensity measured on an 11-point numerical pain rating scale. Pain 2001;94:149–158.

[12] Harden RN, Bruehl S, Perez, R S G M, Birklein F, Marinus J, Maihofner C, Lubenow T, Buvanendran A, Mackey S, Graciosa J, Mogilevski M, Ramsden C, Chont M, Vatine JJ. Validation of proposed diagnostic criteria (the “Budapest Criteria”) for Complex Regional Pain Syndrome. Pain 2010;150:268–274.

[13] Harden RN, Swan M, King A, Costa B, Barthel J. Treatment of complex regional pain syndrome - Functional restoration. Clin J Pain 2006;22:420–424.

[14] Lewis JS, McCabe CS. Body perception disturbance in CRPS. Practical Pain Management 2010;10:60–66.

[15] Lewis JS, Schweinhardt P. Perceptions of the painful body: The relationship between body perception disturbance, pain and tactile discrimination in complex regional pain syndrome. European Journal of Pain 2012;16:1320–1330.

[16] Lewis JS, Coales K, Hall J, McCabe CS. ‘Now you see it, now you do not’: sensory– motor re-education in complex regional pain syndrome. Hand Therapy 2011;16:29–38.

[17] Lewis JS, Kersten P, McCabe CS, McPherson KM, Blake DR. Body perception disturbance: A contribution to pain in complex regional pain syndrome (CRPS). Pain 2007;133:111–119.

[18] Longo MR. Implicit and explicit body representations. Eur Psychol 2015;20:6–15.

[19] Longo MR, Schuur F, Kammers MPM, Tsakiris M, Haggard P. Self awareness and the body image. Acta Psychol 2009;132:166–172.

[20] Longo MR, Haggard P. Implicit body representations and the conscious body image. Acta Psychol 2012;141:164–168.

[21] Maihöfner C, Handwerker HO, Neundörfer B, Birklein F. Patterns of cortical reorganization in complex regional pain syndrome. Neurology 2003;61:1707–1715.

[22] McCormick ZL, Gagnon CM, Caldwell M, Patel J, Kornfeld S, Atchison J, Stanos S, Harden RN, Calisoff R. Short-Term Functional, Emotional, and Pain Outcomes of Patients with Complex Regional Pain Syndrome Treated in a Comprehensive Interdisciplinary Pain Management Program. Pain Med 2015;16:2357–2367.

[23] Melzack R. Phantom limbs. Reg Anesth 1989;14:208–211.

[24] Michal M, Adler J, Reiner I, Wermke A, Ackermann T, Schlereth T, Birklein F. Association of Neglect-Like Symptoms with Anxiety, Somatization, and Depersonalization in Complex Regional Pain Syndrome. Pain Medicine 2017;18:764.

[25] Mölbert SC, Klein L, Thaler A, Mohler BJ, Brozzo C, Martus P, Karnath H, Zipfel S, Giel KE. Depictive and metric body size estimation in anorexia nervosa and bulimia nervosa: A systematic review and meta-analysis. Clinical Psychology Review 2017;57:21–31.

[26] Moseley GL. Distorted body image in complex regional pain syndrome. Neurology 2005;65:773.

[27] Moseley GL, Gallance A, Iannetti GD. Spatially defined modulation of skin temperature and hand ownership of both hands in patients with unilateral complex regional pain syndrome. Brain 2012;135:3676–3686.

[28] Moseley GL, Gallace A, Spence C. Bodily illusions in health and disease: Physiological and clinical perspectives and the concept of a cortical ‘body matrix’. Neuroscience & Biobehavioral Reviews 2012;36:34–46.

[29] Moseley GL. Graded motor imagery is effective for long-standing complex regional pain syndrome: A randomised controlled trial. Pain 2004;108:192–198.

[30] Moseley GL, Flor H. Targeting cortical representations in the treatment of chronic pain: A review. Neurorehabil Neural Repair 2012;26:646–652.

[31] Paillard J. Body schema and body image - A double dissociation in deafferented patients. Motor Control, Today and Tomorrow. 1999:197–214.

[32] Peltz E, Seifert F, Lanz S, Muller R, Maihöfner C. Impaired Hand Size Estimation in CRPS. J Pain 2011;12:1095–1101.

[33] Pitron V, Alsmith A, de Vignemont F. How do the body schema and the body image interact? Consciousness and Cognition 2018;65:352–358.

[34] Pourmand A, Davis S, Marchak A, Whiteside T, Sikka N. Virtual Reality as a Clinical Tool for Pain Management. Curr Pain Headache Rep 2018;22.

[35] Preston C, Newport R. Analgesic effects of multisensory illusions in osteoarthritis. Rheumatology 2011;50:2314–2315.

[36] Schaefer M, Flor H, Heinze H, Rotte M. Morphing the body: Illusory feeling of an elongated arm affects somatosensory homunculus. Neuroimage 2007;36:700–705.

[37] Singh G, Willen SN, Boswell MV, Janata JW, Chelimsky TC. The value of interdisciplinary pain management in complex regional pain syndrome type I: a prospective outcome study. Pain physician 2004;7:203.

[38] Smart KM, Wand BM, O’Connell NE. Physiotherapy for pain and disability in adults with complex regional pain syndrome (CRPS) types I and II. Cochrane Database Syst Rev 2016;2016.

[39] Tracey I, Mantyh PW. The Cerebral Signature for Pain Perception and Its Modulation. Neuron 2007;55:377–391.

[40] Turner-Stokes L, Goebel A, Guideline Dev Grp. Complex regional pain syndrome in adults: concise guidance. Clin Med 2011;11:596–600.

